# Analysis of Drug Formulary Exclusions from the Patient’s Perspective: 2023 Update

**DOI:** 10.1101/2023.11.01.23297921

**Authors:** Sara Chea, Anne M. Sydor, Robert Popovian

**Author notes:** **CORRESPONDING SENIOR AUTHOR:**: Robert Popovian. **COMPETING INTERESTS DISCLOSURE:** Robert Popovian is the founder of Conquest Advisors, LLC, owns stocks of biopharmaceutical companies, and was an employee of Pfizer for over two decades. He is a member of the Board of Councilors, University of Southern California, School of Pharmacy, Board of Directors, for University Pharmaco, LLC, Adjunct Clinical Faculty at Rutgers University, School of Pharmacy and serves as a consultant for the biopharmaceutical industry. The authors declare no other relevant conflicts of interest or financial relationships. **FUNDING:** This study was funded by unrestricted educational grants from Amgen.

## Abstract

**Objective:** Pharmacy benefit management companies (PBMs) often determine medication reimbursement, out-of-pocket costs, and access through formularies. Formularies were initially intended to ensure the use of cost-effective medication. Today, formularies are designed to maximize concessions (i.e., rebates, discounts, fees, and other concessions) to PBMs from the biopharmaceutical industry. Formulary exclusions enhance the ability to drive profits through rebate contracting for PBMs. Our 2022 research analyzed whether formulary exclusions benefit patients medically or economically. This update provides an analysis of exclusions based on the 2023 Express Scripts (ESI) national formulary.

**Methods:** We analyzed ESI’s 2023 national preferred formulary exclusions. ESI is the second-largest PBM in the U.S. and makes its national preferred formulary exclusions list publicly available. We categorized substitutions as equivalent (same active agent used) vs. therapeutic (different active agent). From a patient perspective, we evaluated each exclusion by potential clinical or economic outcomes and compared it to the results from the 2022 analysis.

**Results:** More than half (57.4%) of the formulary exclusions had questionable economic or medical benefits or both for patients. The results demonstrate a 9% increase in questionable patient benefits compared with 48.4% in 2022.

**Conclusions:** Because patient co-pays and deductibles are based on retail prices, some formulary exclusions force patients to pay substantially more for a preferred drug or use a medication with questionable medical benefits for their condition. Exclusions also force prescribers to choose treatments that may have adverse financial or medical outcomes for their patients.

## INTRODUCTION

The United States healthcare marketplace consists of consumers covered by insurance companies providing prescription medication biopharmaceutical benefits. The insurance plan may be sponsored by the federal or state government, individual consumers, employers, and others who purchase health insurance. Biopharmaceutical benefits are managed by pharmacy benefit management companies (PBM) on behalf of plan sponsors.

Three of the largest PBMs, CVS Health (Caremark), Cigna (Express Scripts and Ascent Health Services), and United Health (OptumRx), process more than 79% of retail prescriptions.^1^ The three largest PBMs also control 65% of the prescription drug revenue from specialty medicines in the U.S.^2^ Specialty medicines are primarily brand-name medicines, providing PBMs with significant revenue through rebate contracting. Over the last several years, the three PBMs have been acquired by insurance companies.^3^ The horizontal integration of insurance companies and PBMs has created a massive healthcare delivery monopoly concerning drug coverage.

PBMs construct formularies, which are lists of medications that are covered through a patient pharmacy benefit. Formularies typically favor the use of the most cost-effective medicine, which was the original intent of developing formularies.^4^ Tiered formularies can increase medication availability by allowing patients access to all drugs if a patient is willing to pay a fixed out-of-pocket cost based on the preferred tier.^5^ Formularies are subject to change annually, and changes are made based on FDA approval, clinical data, or contractual agreements with individual biopharmaceutical companies.

Today, however, the purpose of formularies has shifted to increase the use of medicines that maximize rebates, discounts, fees, and other concessions from biopharmaceutical companies.^6-7^ Over the past several years, as confirmed by Texas Department of Health Insurance data, PBMs pass less than 1% of rebates they collect back to patients.^8^

PBMs now release annual exclusion lists or lists of medicines that will not be covered through the PBM under any circumstance. The exclusions are used as leverage to gain more rebates and fees from the biopharmaceutical industry.^9^ The number of exclusions has increased by an average of 34% each year from 2014-2022.^10-11^ The practice of growing formulary exclusions limits patient drug accessibility. It is essential to note that if a drug is excluded, a patient would have to pay the entire cost out-of-pocket for that medication, which most patients cannot afford. Finally, excluding medicines from formularies creates an environment where a patient whose disease is stable with a particular treatment may be forced to switch therapies, possibly jeopardizing a patient’s well-being. For example, a study of 775 patients whose cardiovascular drugs were excluded found a 51% increase in outpatient emergency department visits within six months of patients being switched to a different medication.^12^

Multiple analyses have quantified the number of exclusions and the types of medicines. The research by the Global Healthy Living Foundation (GHLF) of the Express Scripts (ESI) National Preferred Formulary in 2022 was the first to evaluate whether exclusions were clinically or financially beneficial from a patient’s perspective.^13^ The research concluded that almost half of the 563 excluded medications that were not generic equivalents or insulin devices had questionable economic or medical benefits for patients.

### Purpose of the Study

The current study provides an update based on the 2023 Express Scripts (ESI) 2023 National Preferred formulary.^14^ Similar to the previous study, we evaluated all listed exclusions for their impact on patient’s clinical and financial outcomes.^13^ We chose ESI because it is the second largest PBM in the United States and is a publicly available national preferred formulary exclusion list annually. We compared the exclusions from 2022 and 2023 to identify changes and trends.

## METHODS

We first categorized each exclusion as an equivalent substitution, a therapeutic substitution, or excluded without an alternative substitution.

### Equivalent Substitutions

Equivalent substitutions include:

- Brand, generic, or biosimilar medicine is excluded in favor of a preferred generic or biosimilar medication containing the same active ingredient,
- Brand medicine is excluded in favor of another brand medicine that has the same active ingredient,
- Generic or biosimilar medicine is excluded in favor of a brand-name medicine containing the same active ingredient
- Brand, biosimilar, or generic medicine is excluded in favor of a different formulation with the same active ingredient. Formulation differences included different dosages, routes of administration, combination pills, and drug delivery systems.

### Therapeutic Substitutions

Therapeutic substitutions include:

- Brand, biosimilar, or generic medicine is excluded in favor of another brand or generic drug that does not contain the same active ingredient.

### Therapeutic Substitutions

Therapeutic substitutions included:

- Brand, biosimilar, or generic medicine is excluded without any alternative recommended by the formulary.

### Clinical and Economic Outcomes

We then categorized each exclusion based on the potential clinical or economic outcome from a patient perspective.

A clearly positive clinical outcome was defined as:

- The formulary includes the medicine with the same active ingredient and formulation in the form of a brand name, generic, or biosimilar medicine as an excluded drug.

A clearly positive economic outcome was defined as:

- An alternative to an excluded drug is an equivalent substitution with a generic or biosimilar with a similar active ingredient and formulation and thus is presumed cheaper than the excluded brand medication.

Exclusions that did not meet the above criteria were considered to have questionable medical and/or economic benefits. This is because the medical outcome of a therapeutic substitution may, but will not necessarily, cause new side effects or worsening of disease. A medical benefit to the patient for a biopharmaceutical other than what was prescribed cannot be recognized without knowing why the particular excluded drug was prescribed. Similarly, the economic effect of a substitution that is not a generic equivalent substitution cannot be determined since insurance companies, PBMs, and biopharmaceutical companies do not openly share the net prices paid for individual medications.

## Results

In the ESI formulary, the total exclusions increased by 125 from 2022 to 2023 (Table 1), a 22.2% (125/563) increase year over year. Exclusions with no preferred alternative grew at the fastest rate, more than doubling from 9 to 20. In 2022 and 2023, non-equivalent substitutions accounted for one-third of all the exclusions.

**Table 1:** Total Number of Exclusions and Modifications from 2022 to 2023.

| **Table 1: Total Number of Exclusions and Modifications from 2022 to 2023.** |  |  |  |  |
| --- | --- | --- | --- | --- |
| **Substitution** | **2023 Number** | **2022 Number** | **Increase** | **% Increase** |
| Equivalent substitutions | 479 | 386 | 93 | 24.1% |
| Therapeutic substitutions | 189 | 168 | 21 | 12.5% |
| No substitutions | 20 | 9 | 11 | 122% |
| **Total Exclusions** | **688** | **563** | **125** | **22.2%** |

Formulation substitutions grew most quickly, more than doubling year over year from 58 to 126 (Table 2). Generic/biosimilar substitution with a different generic/biosimilar alternative formulation grew at the fastest rate, tripling from 12 to 36 (Table 3). There was also an increase from 41 to 49 equivalent substitutions for which a brand-name drug was preferred (Tables 2 and 3). The number substitutions that favored a brand-name drug over a generic or biosimilar increased from 10 in 2022 to 13 in 2023.

**Table 2:** Equivalent Substitutions Categorized by Class Excluded and Substituted.

|  |  |  |  |  |  |
| --- | --- | --- | --- | --- | --- |
| **Excluded** | **Preferred alternative** | **2023**<br>**Number** | **2022**<br>**Number** | **Increase** | **%**<br>**Increase** |
| Brand-name | Generic or biosimilar | 293 | 273 | 20 | 7.3% |
|  | Brand-name | 32 | 30 | 2 | 6.0% |
| Generic or biosimilar | Generic or biosimilar | 21 | 20 | 1 | 5.0% |
|  | Brand-name | 7 | 5 | 2 | 40.0% |
| Formulation substitutions |  | 126 | 58 | 68 | 117% |
| **Total Equivalent Substitutions** |  | **479** | **386** | **93** | **24.1%** |

**Table 3:** Formulation Substitutions Categorized by Class Excluded and Substituted.

|  |  |  |  |  |  |
| --- | --- | --- | --- | --- | --- |
| **Excluded** | **Preferred Alternative** | **2023**<br>**Number** | **2022**<br>**Number** | **Number**<br>**Change** | **%**<br>**Increase** |
| Brand-name | Generic or biosimilar | 80 | 40 | 40 | 100% |
|  | Brand-name | 9 | 6 | 3 | 50% |
| Generic or biosimilar | Generic or biosimilar | 36 | 12 | 24 | 200% |
|  | Brand-name | 1 | 0 | 1 | 0.8% |
| **Total Formulation Substitutions** |  | **126** | **58** | **68** | **117%** |

Therapeutic substitutions increased from 168 in 2022 to 189 in 2023 (Table 4). Therapeutic substitutions excluding brand name drugs for generic drugs increased the most, from 84 to 100, in 2023. Brand name to brand name and generic to generic therapeutic substitutions were similar from 2022 to 2023.

**Table 4:** Therapeutic Substitutions Categorized by Class Excluded and Substituted.

|  |  |  |  |  |  |
| --- | --- | --- | --- | --- | --- |
| **Excluded** | **Preferred Alternative** | **2023**<br>**Number** | **2022**<br>**Number** | **Increase** | **%**<br>**Increase** |
| Brand-name | Generic or biosimilar | 100 | 84 | 16 | 19% |
|  | Brand-name | 70 | 67 | 3 | 4.4% |
| Generic or biosimilar | Generic or biosimilar | 13 | 12 | 1 | 8.3% |
|  | Brand-name | 6 | 5 | 1 | 20.0% |
| **Total Therapeutic Substitutions** |  | **189** | **168** | **21** | **12.5%** |

Lastly, Table 5 lists the economic and medical benefits of exclusions from the patients’ perspective. The number of exclusions that gave patients economic benefits stayed the same at 293 exclusions. The total number of exclusions with questionable benefits was 395 out of 688 (57.4%), a 9.6% increase from 2022 when exclusions with questionable benefits accounted for 47.9% of all exclusions.

**Table 5:**
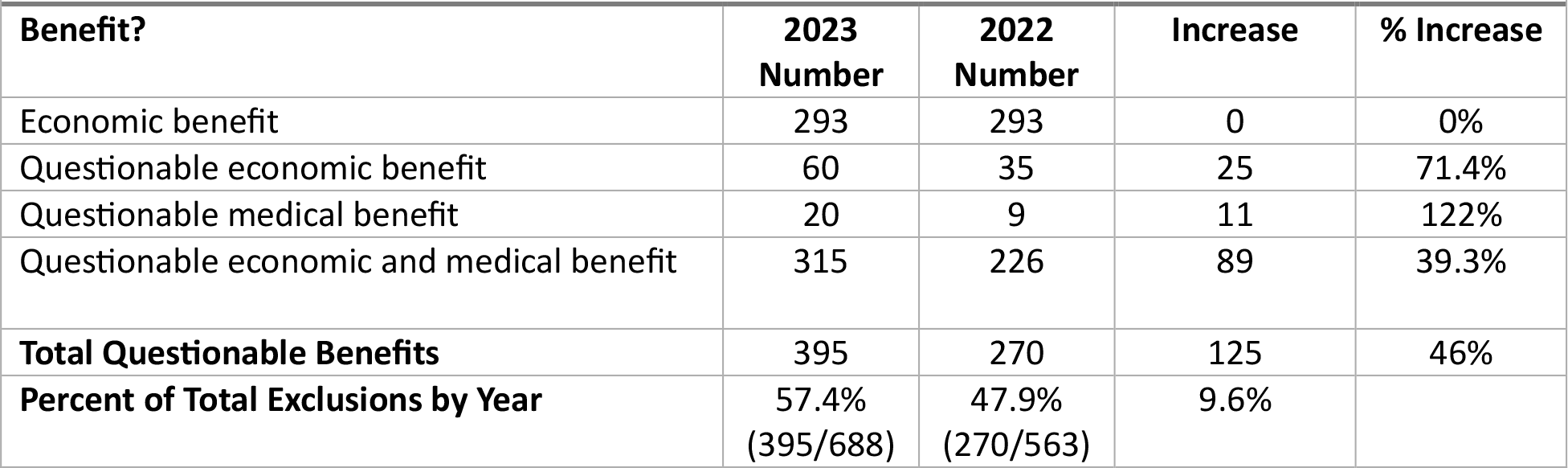
Economic and Medical Benefits of Exclusion to Patient.

| **Table 5: Economic and Medical Benefits of Exclusion to Patient.** |  |  |  |  |
| --- | --- | --- | --- | --- |
| **Benefit?** | **2023**<br>**Number** | **2022**<br>**Number** | **Increase** | **% Increase** |
| Economic benefit | 293 | 293 | 0 | 0% |
| Questionable economic benefit | 60 | 35 | 25 | 71.4% |
| Questionable medical benefit | 20 | 9 | 11 | 122% |
| Questionable economic and medical benefit | 315 | 226 | 89 | 39.3% |
| **Total Questionable Benefits** | **395** | **270** | **125** | **46%** |
| **Percent of Total Exclusions by Year** | **57.4%**<br>**(395/688)** | **47.9%**<br>**(270/563)** | **9.6%** |  |

### Limitations

The study conducted was representative of a single plan year of formulary exclusions compared across two years. However, data supports that exclusions with questionable economic and medical benefits are increasing annually. Because other formularies are not publicly available, it is not possible to determine if this is truly representative of the formulary exclusion practices of all PBMs. The analysis is based on a national exclusionary formulary. However, plan sponsors may adopt or adjust the formulary based on their needs. As discussed, beneficial vs. questionable economic outcomes could only be assumed because the actual price paid by an insurer or PBM to a biopharmaceutical company for any medication is not publicly disclosed. Lastly, the analysis has a level of subjectiveness in categorizing every exclusion and its alternative as a questionable economic or medical benefit or both. However, for every point of decision analysis, the perspective of the patient’s access to the most appropriate drug for their disease state was considered of utmost importance.

## Discussion

The original intent of a formulary was to prioritize the use of cost-effective medicines. However, today, formularies are mechanisms to maximize rebates to PBMs. The number of exclusions for national ESI formulary has increased by 22.2% from 2022 to 2023 (Table 1) and 57.4% of these have questionable medical or economic benefits to patients (Table 5), an increase of almost 10% compared to 2022. As such, patients are forced to accept therapies that may not be medically or economically beneficial to them. Our study also demonstrates that all new exclusions are of questionable economic or medical benefit to the patient since the number of financially beneficial exclusions remained the same from 2022 to 2023 (n=293). Exclusions now pose a barrier to medication access and favorable treatment outcomes and are a growing threat to patients’ medication access.

In some cases, the ESI formulary excludes medicines without providing any alternatives to patients and healthcare professionals, potentially forcing patients to forgo medically necessary treatments. In 2023, the number of exclusions without an option doubled, from 9 to 20 exclusions, a 122% increase. An example of such exclusion is ONUREG (azacytidine), a brand-name oncology drug prescribed for the rare indication of acute myeloid leukemia. Interestingly, options for ONUREG were listed in the 2022 ESI formulary exclusions list [15], but alternatives were removed in 2023. In all 20 cases containing no substitutions, the exclusions are the only disease-modifying treatments available for the condition treated by the excluded medication.

Exclusions due to formulation differences may adversely affect patient outcomes due to medication non-adherence. For example, different formulations, such as combination pills, or user-friendly delivery mechanisms can increase medication adherence, improving patient care while reducing healthcare costs. Within the equivalent substitution category, there was a 117% increase in formulation substitutions from 58 in 2022 to 126 in 2023 (Table 2). It is essential to note that 126 of equivalent substitutions may be deemed therapeutic substitution because the excluded formulation is not the same as the alternatives covered by the PBM.

In 13 cases, the exclusions favor brand medicines that are significantly more expensive than the excluded generics or authorized generics. (Table 3 and 4). For example, zomitriptan, a generic nasal spray for migraine treatment, is excluded in preference for Zomig nasal brand name medicine. In another case, ESI has excluded Amjevita with a National Drug Code (NDC) starting with 72511 and instead covers Amjevita with an NDC starting with 55513.^14^ Amjevita is a biosimilar of the brand name Humira. The Amjevita manufacturer Amgen has offered two price points, one at a 5% discount to the Humira (NDC code 55513) listed price and one at a 55% discount (NDC code 72511).^16^ Such practices affirm that formularies can be used as a rebate profitability tool for PBMs since they prefer higher-priced and more highly rebated drugs instead of lower-cost generic, authorized generic, or biosimilar alternatives. Opaque contracts between PBMs and biopharmaceutical companies veil these hidden excess costs. The rebates, fees, and other concessions gained by PBMs from the biopharmaceutical industry do not necessarily translate as cost savings to plan sponsors such as the government, employers, and individual patients. In addition, patients also take on the burden of out-of-pocket deductibles based on the retail prices of the formulary drug, which can be at a more significant cost than the excluded ones. In the past 10 years, exclusion lists from the three largest PBMs in the nation have continued to expand exponentially, reflecting the exponential growth of patients affected by such exclusions.

## Conclusions

Formulary exclusions have become the norm in managing drug benefits by PBMs. They have been growing in number year over year. Although some formulary exclusions may be clinically and economically justified, a significant number require healthcare professionals to make medical decisions that may not be in the patient’s best interest or aligned with current standards of care. Uniformly, such practices continue to blur the line between insurance coverage and medical practice, highlighting the need to reform the drug rebating system.

## Data Availability

All data produced in the present study are available upon reasonable request to the authors

## References

1. Fein AJ (2023) The Top Pharmacy Benefit Managers of 2022: Market Share and Trends for the Biggest Companies. Drug Channels.

2. Fein AJ (2023) DCI’s Top 15 Specialty Pharmacies of 2022: Five Key Trends About Today’s Marketplace. Drug Channels.

3. Richman B, Schulman K (2018) Mergers between health insurers and pharmacy benefit managers could be bad for your health. STAT 2022.

4. Kelly D A (2018) A brief history of drug formularies and what to expect in 2019. CoverMyMeds. 2018. Accessed.

5. Werble C (2017) Health care policy brief: formularies. Health Affairs Accessed April 18, 2022.

6. Alston M, Dieguez G, Tomicki S (2018) A primer on prescription drug rebates: Insights into why rebates are a target for reducing prices. Milliman.

7. Fein AJ (2018) Janssen’s new transparency report: a peek behind the drug pricing curtain raises troubling questions about rebates. Drug Channels.

8. Texas Department of Health Insurance, Life, Health and Accident Insurance Reports, Prescription Drug Cost Transparency, Pharmacy Benefit Managers, https://www.tdi.texas.gov/reports/report3.html

9. Avalere (2015) New analysis from Avalere Health finds that some exchange plans place all drugs used to treat complex diseases –such as HIV, cancer, and multiple sclerosis - on the highest drug formulary cost-sharing tier. Avalere Analysis.

10. Fein AJ (2023) The Big Three PBMs’ 2023 Formulary Exclusions: Observations on Insulin, Humira, and Biosimilars. Drug Channels.

11. Xcenda (2022) Skyrocketing growth in PBM formulary exclusions raises concerns about patient access. AmericaSource Bergen: Xcenda.

12. Xcenda (2023). Issue brief: Assessing the impact of formulary exclusion on healthcare costs and outcomes for patients on therapy for certain chronic conditions. AmericaSource Bergen: Xcenda.

13. Popovian R, Sydor AM, Pits P (2022) Analysis of the Drug Formulary Exclusions from a Patient’s Perspective. Health Sci J. Vol. 16 No. 3:937.

14. 2023 National Preferred Formulary Exclusions. Express Scripts. https://cut.ly/lwjIAHmQ

15. 2022 National Preferred Formulary Exclusions. Express Scripts.

16. Becker, Z (2023). Amgen’s Humira Biosimilar Amjevita hits the market with 2 different list prices. Fierce Pharma.

